# Transmission of COVID-19 in 282 clusters in Catalonia, Spain: a cohort study

**DOI:** 10.1101/2020.10.27.20220277

**Authors:** Michael Marks, Pere Millat-Martinez, Dan Ouchi, Chrissy h. Roberts, Andrea Alemany, Marc Corbacho-Monné, Maria Ubals, Aurelio Tobias, Cristian Tebé, Ester Ballana, Martí Vall-Mayans, Camila G-Beiras, Nuria Prat, Jordi Ara, Bonaventura Clotet, Oriol Mitjà

## Abstract

**Background:** There remains limited data on what variables affect risk of transmission of SARS-CoV-2 and developing symptomatic Covid-19 and in particular the relationship to viral load (VL). We analysed data from linked index cases and their contacts to explore factors associated with transmission of SARS-CoV-2.

**Methods:** Patients were recruited as part of a randomized control trial, conducted between March to April 2020, that aimed to assess if hydroxychloroquine reduced transmission of SARS-CoV-2. Non-hospitalised Covid-19 cases and their contacts were identified through the local surveillance system. VL, measured by quantitative PCR from a nasopharyngeal swab, was assessed at enrollment, at day 14, and whenever the participant reported Covid-19-like symptoms. Risk of transmission, developing symptomatic disease and incubation dynamics were evaluated using regression analysis.

**Findings:** We identified 314 cases, 282 of which had at least one contact (753 contacts in total). Ninety (33%) of 282 clusters had at least one transmission event. The secondary attack rate was 16% (125/753), with a variation from 12% to 24% for VL of the index case of <10^6^, and >10^9^ copies/mL, respectively (OR per log_10_ increase in VL 1.3 95%CI 1.1–1.6). Increased risk of transmission was also associated with household contact (OR 2.7; 1.4–5.06) and age of the contact (OR 1.02 per year; 1.01–1.04). The proportion of PCR positive contacts who developed symptomatic Covid-19 was 40.3% (181/449), with a variation from 25% to 60% for VL of the contact <10^7^, and >10^9^ copies/mL (HR log_10_ increase in VL 1.12; 95% CI 1.05 – 1.2). Time to onset of symptomatic disease decreased from a median of 7 days (IQR 5–10) for individuals with an initial viral load <10^7^ to 6 days (4–8) and 5 days (3–8) for individuals with an initial viral load of 10^7^–10^9^ and >10^9^, respectively.

**Interpretation:** Viral load of index cases is a leading driver of SARS-CoV-2 transmission. The risk of symptomatic Covid-19 is strongly associated with viral load of contacts at baseline and shortens the incubation time in a dose-dependent manner.

**Funding:** Crowdfunding campaign YoMeCorono (http://www.yomecorono.com/), and Generalitat de Catalunya. Support for laboratory equipment from Foundation Dormeur.

**Research in context:** *Evidence before this study:* In September 2020, we searched PubMed database for articles reporting on factors influencing transmission and the risk of developing symptomatic disease. Search terms included “Covid-19”, “SARS-CoV-2”, “transmission”, “incubation time”, and “risk”, with no language restrictions. By 20^th^ September, various authors had reported on retrospective analyses of clusters of index cases and their corresponding contacts, as well as series of patients who developed symptomatic Covid-19 disease after PCR positive result. Besides describing the secondary attack rate, various authors identified risk factors for transmission associated with the place and duration of exposure and the lack of use of personal protective equipment. A single study suggested that symptomatic individuals might be more likely to transmit than asymptomatic cases but we found no clear evidence regarding the influence of viral load of the index case on transmission risk. Similarly, although various retrospective series of patients with positive PCR results had reported incubation times elsewhere, the characteristics of index case and contacts that may influence the risk of developing symptomatic Covid-19 and the time to this event had been barely addressed.

*Added value of this study:* We analyzed data from a large cluster-randomized clinical trial on post-exposure therapy for Covid-19 that provide new information on SARS-CoV-2 transmission dynamics. Several design components add value to this dataset. Notably, quantitative PCR was available for the index cases to estimate risk of transmission. Furthermore, quantitative PCR was also performed on asymptomatic contacts at the time of enrollment allowing to investigate the dynamics of symptomatic disease onset among them. We found that the viral load of the index case was the leading determinant of the risk of SARS-CoV-2 PCR positivity among contacts. Among contacts who were SARS-CoV-2 PCR positive at baseline, viral load significantly influenced the risk of developing the symptomatic disease in a dose-dependent manner. This influence also became apparent in the incubation time, which shortened with increasing baseline viral loads.

*Implication of all the available evidence:* Our results provide important insights into the knowledge regarding the risk of SARS-CoV-2 transmission and Covid-19 development. The fact that the transmission risk is primarily driven by the viral load of index cases, more than other factors such as their symptoms or age, suggests that all cases should be considered potential transmitters irrespective of their presentation and encourages assessing viral load in cases with a larger number of close contacts. Similarly, our results regarding the risk and expected time to developing symptomatic Covid-19 encourage risk stratification of newly diagnosed SARS-CoV-2 infections based on the initial viral load.

## INTRODUCTION

According to current evidence, Covid-19 is primarily transmitted from person to person through respiratory droplets, as well as indirect contact, through transfer of the virus from contaminated fomites to the mouth, nose, or eyes.^1,2^ As with most respiratory viral infections there is likely to be some contribution from smaller aerosols but their relative contribution compared to droplets remains unclear. Several outbreak investigation reports have shown that Covid-19 transmission can be particularly effective in confined indoor spaces such as workplaces including factories, churches, restaurants, shopping centers, or healthcare settings.^3–6^ In Spain, and many other countries, healthcare workers have experienced a high rate of Covid-19 infection.^7^

The availability of data regarding the factors that may enhance transmission is essential for designing interventions to control SARS-CoV-2 spread. Currently available data provide information on the risk of transmission related to the place and duration of exposure, and the use of respiratory and eye protection^1,3– 5,8^ but not on other factors related to the characteristics of index cases and their contacts. Over the course of infection, the virus has been identified in respiratory tract specimens 1–2 days before the onset of symptoms, and it can persist for prolonged periods over several weeks after the onset of symptoms in mild cases.^9^ However, the detection of viral RNA by PCR does not necessarily equate with infectivity, and the exact relationship between viral load and risk of transmission from a case is still not clear.^10,11^ Studies investigating case-contact pairs have reported highly variable secondary attack rates (i.e., range 0.7% to 75%), depending on the type of exposure-duration, place, pre- or post-symptomatic.^12–15^

Another challenge for public health interventions is the risk stratification of infected individuals for developing symptomatic illness. On the other hand, a living systematic review estimated that the proportion of PCR-positive infected contacts that progress to symptomatic disease is approximately 70-80%.^16,17^ Estimates of mean or median incubation period have been consistently between 5–7 days.^18–20^ Whilst there has been a suggestion that viral load of cases may potentially be associated with risk of disease or transmission there is currently no published data directly addressing this question and little is known about factors that may contribute to variation on the risk of developing Covid-19 symptoms or the incubation periods among infected individuals.

The overall aim of this study was to evaluate transmission dynamics of SARS-CoV-2 in the context of a trial of post-exposure prophylaxis. Specifically, the objectives of the study were threefold: (a) to investigate the association between clinical and demographic features of cases and viral load, (b) to evaluate the effect of viral load on SARS-CoV-2 transmission to close contacts, and (c) to determine the influence of viral load in the exposed on development of symptoms and on the incubation period.

## METHODS

### Study design

This was a post-hoc analysis of data collected in the BCN PEP CoV-2 Study (NCT04304053), a cluster-randomized trial that included PCR-confirmed Covid-19 cases and their close contacts. The trial occurred between Mar 17 to Apr 28, 2020, during the SARS-CoV-2 outbreak, in three out of nine healthcare areas in Catalonia (North-East Spain): *Catalunya central, Àmbit Metropolità Nord, and Barcelona Ciutat*, total target population 4,206,440 people. The study protocol of the BCN PEP CoV-2 Study was approved by the ethics committee of Hospital Germans Trias Pujol, (Badalona, Spain). Written informed consent was obtained from all participants. Full details of the original study are reported elsewhere.^21^

Covid-19 cases were identified using the electronic registry of the Epidemiological Surveillance Emergency Service of Catalonia (SUVEC) of the Department of Health.^22^ Following government ordinance, the SUVEC registered all new Covid-19 diagnoses occurred from March 16, 2020. The surveillance system included active tracing of all contacts with recent history of exposure, defined as being in contact with a SARS-CoV-2 PCR positive case during more than 15 minutes within two meters.

All Covid-19 cases included in the present analysis were non-hospitalized adults (i.e., ≥ 18 years of age) with quantitative PCR result available at baseline, mild symptom onset within five days before enrollment, and no reported symptoms of SARS-CoV-2 infections in their accommodation (i.e., household or nursing home) or workplace within the 14 days before enrollment. Contacts selected for the analysis were adults with a recent history of exposure and absence of Covid-19-like symptoms within the seven days preceding enrolment. Contacts were exposed to the index case as either a healthcare worker, a household contact, a nursing home worker, or a nursing home resident.

### Study procedures and data collection

A dedicated outbreak field team visited cases and contacts at home or nursing home on days 1 (enrollment) and 14. At the first clinical assessment on day 1 they conducted a baseline assessment, including a questionnaire for symptoms of Covid-19 and collected relevant epidemiological information using a structured interview: time of first exposure to the index case, place of contact (hospital, home, nursing care facility), routine use of a mask of both when in close proximity to the index case, the case and the contact, and sleep location concerning the index case (e.g., same room, same house). Symptoms surveillance consisted of active monitoring by phone on days 3, and 7, a home visit on day 14, and passive monitoring whenever the participants developed symptoms. Participants who developed symptoms were visited the same day they notified symptom onset (unscheduled visits) by the field team, which recorded the date of symptom onset, type of symptoms from a pre-specified checklist, and symptom severity, graded on a 1-to-4 scale.

Serial SARS-CoV-2 PCR test and viral load titration on nasopharyngeal swab were conducted on day 1 and day 14 to all participants, and on any unscheduled visit when the participant notified the onset of Covid-19 symptoms. The detection of the SARS-CoV-2 virus was performed from nasopharyngeal swabs at SYNLAB Diagnostics (Barcelona, Spain) by PCR using TaqMan™ 2019-nCoV Assay Kit according to the manufacturer’s protocol (Catalog number: A47532, Thermo Fischer Scientific Inc.). Viral load was quantified from nasopharyngeal swabs at IrsiCaixa laboratory (Badalona, Spain) by PCR amplification, based on the 2019-Novel Coronavirus Real-Time RT-PCR Diagnostic Panel guidelines and protocol developed by the American Center for Disease Control and Prevention (CDC).^23^ For absolute quantification, a standard curve was built using 1/5 serial dilutions of a SARS-CoV2 plasmid (2019-nCoV_N_Positive Control, catalog no. 10006625, 2×10^5^ copies/μL, Integrated DNA Technologies) and run in parallel to all PCR determinations.

### Outcomes and definitions

Transmission was characterized by examining the number of infected and uninfected individuals among close contacts to an index case. We defined transmission events as PCR-positivity at any time point (i.e., days 1, 14, or at any other unscheduled PCR testing when participants referred symptoms) of a contact in the same household or workplace within the 14 days following enrollment. We defined the secondary attack rate of viral transmission as the ratio of PCR-positive individuals among close contacts, according to the WHO guidelines.

Development to symptomatic disease was defined as presence of at least one of the following symptoms: fever, cough, difficulty breathing, myalgia, headache, sore throat, new olfactory and taste disorder(s), or diarrhea) and a positive SARS-CoV-2 RT-PCR test. The incubation period was defined as time from first exposure to symptom onset, with later confirmation of infection by PCR.^24^ The earliest possible exposure with the symptomatic index case was determined for each contact individually.

### Study Participants

We selected all eligible individuals within the original trial population for each of the three analyses conducted in the current study. As in the original trial there was no evidence of an impact of hydroxychloroquine on either transmission or development of symptomatic disease we included individuals in both arms of the trial in the current study. Firstly, all Covid-19 cases with quantitative PCR data were included in an analysis of the association between clinical and demographic features of cases and viral load. Secondly, we identified factors associated with transmission using all clusters of an index case (i.e., a Covid-19 case with at least one close contact) and their corresponding contacts for which quantitative viral load was available for the index case. Finally, we assessed the risk of developing symptomatic disease and the variation in the incubation period amongst all contacts with a positive PCR result at baseline, irrespective of available data of their index case.

### Statistical Analysis

We used log transformed viral loads which were approximately normally distributed and which also align with common reporting norms. The relationship between characteristics of cases and viral load was assessed using linear regression considering age (in years), sex, the number of days from reported symptom onset and the presence of absence of five key clinical features namely fever, cough, shortness of breath or rhinitis and anosmia. To identify risk factors for transmission, we used logistic regression model for the risk of transmission utilizing a random-effect model to allow for within cluster variation in the risk of transmission. Factors with potential influence on the risk of transmission included characteristics of the potential transmitter (i.e., age, sex, viral load, and the presence or absence of respiratory symptoms) and contacts (i.e., age, sex, and the type of contact they had with the index case). Finally the risk of developing symptomatic Covid-19 was assessed by fitting a cox-regression model considering the age (in years) and sex of the individual, the presence or absence of cardiovascular disease, chronic respiratory disease and the initial viral load in relation to the time to development of symptomatic disease. Data at 14 days after the first study visit were censored, in line with the follow-up conducted in the original trial. All analyses were conducted in R version 4.0.

## RESULTS

### Sample characteristics

During the investigation period, we identified 314 cases in whom the viral load was tested. Overall, 220 (70.0%) were female and the median age was 41 (IQR 31-52). Of them, 282 had at least one close contact, resulting in the corresponding clusters, with a total of 753 contacts. Clusters had a median of 2 contacts (IQR 1-3) and a maximum of 19 contacts. Most index cases of the clusters were female (n= 202, 71.6%), with an average age of 42 years (SD 13 years) (Table 1).

**Table 1:** Baseline Characteristics of linked transmission clusters.

**Baseline Characteristics of linked transmission clusters**
| Variable |  | Value |
| --- | --- | --- |
| Cluster Size | Median (IQR) | 2 (1-3) |
| Index Case Age | Years – Mean (SD) | 42 (13) |
| Index Case Sex | Female (%) | 202 (64.%) |
| Index Case Log Viral Load | Mean (SD) | 8 (1.8) |
| Contacts Age | Years – Mean (SD) | 42 (15) |
| Contacts Gender | Female (%) | 385 (51.1%) |
|  | Male (%) | 305 (40.5%) |
|  | Missing (%) | 63 (8.4%) |
| Baseline PCR of Contact Case | Positive | 93 (14.2%) |
| Contact | HCW | 254 (33.7%) |
|  | Household | 382 (50.7%) |
|  | Nursing Home | 21 (2.8%) |
|  | Unknown | 96 (12.7%) |

### Index case viral load

The first study visit was performed a median of 4 days (IQR 3-5) after symptom onset. At the first study visit, the mean viral load amongst Covid-19 cases was 10^8^ (10^1.8^). In multivariable linear regression the viral load amongst cases was higher in individuals who reported fever (Table 2) and negatively associated with the presence of anosmia but there was no association between the age or sex of the Covid-19 case nor the presence of reported dyspnea or cough. As anticipated viral load was negatively associated with the number of days since symptom onset.

**Table 2:** Univariable and multivariable linear regression of association between Index case variables and log_10_ viral load.

| Characteristic | | Log<sub>10</sub> Viral Load/ml | Unadjusted $\beta$ coefficient (95% CI) | p | Adjusted $\beta$ coefficient (95% CI) | p |
| --- | --- | --- | --- | --- | --- | --- |
| Case Age |  | N/A | 0.002 (-0.02 – 0.02) | 0.78 | 0.008 (-0.01 – 0.02) | 0.38 |
| Case Sex | Male | 8.15 (7.54 – 8.77) | Reference |  | Reference |  |
|  | Female | 8.04 (7.47 – 8.6) | -0.238 (-0.72 – 2.4) | 0.33 | -0.22 (-0.61 – 0.34) | 0.59 |
| Days from Symptom Onset |  | NA | -0.17 (-0.26 – 0.0.8) | 0.0002 | -0.16 (-0.24 – 0.07) | 0.0004 |
| Cough | Absent | 7.82 (7.24 – 8.41) | Reference |  | Reference |  |
|  | Present | 8.37 (7.78 – 8.95) | 0.66 (0.22 – 1.1) | 0.003 | 0.41 (-0.02 – 0.84) | 0.06 |
| Dyspnea | Absent | 7.97 (7.5-8.43) | Reference |  | Reference |  |
|  | Present | 8.22 (7.45-8.99) | 0.27 (-0.40 – 0.94) | 0.42 | 0.28 (-0.35 – 0.92) | 0.38 |
| Fever | Absent | 7.77 (7.16 – 8.38) | Reference |  | Reference |  |
|  | Present | 8.42 (7.86-8.98) | 0.80 (0.36 – 1.24) | 0.0004 | 0.43 (0.00 – 0.87) | 0.05 |
| Anosmia | Absent | 8.32 (7.76 – 8.88) | Reference |  | Reference |  |
|  | Present | 7.87 (7.25-8.49) | -0.57 (-1.0 - -0.09) | 0.02 | -0.54 (-1.0 – 0.09) | 0.02 |
| Rhinitis | Absent | 7.60 (7.23 – 7.98) | Reference |  | Reference |  |
|  | Present | 8.59 (7.65-9.52) | 0.88 (-0.05 – 1.82) | 0.06 | 0.77 (-0.11 – 1.66) | 0.09 |

### Cluster-level transmission

For our risk factor analysis on SARS-CoV-2 transmission we used linked case and contact data of 282 clusters with 753 contacts. At the cluster level, 90 (33.3%) of the 282 clusters had at least one transmission event, with a highly skewed distribution of the number of transmission events per cluster (Figure 1A). The first visit for contacts took place a median of 5 days (IQR 4-7 days) after their first possible exposure to the index case. A total of 125 (16.6%) of 753 contacts had a PCR positive result over the study period. The proportion of contacts who tested positive for SARS-CoV-2 within a cluster (secondary attack rate) progressively increased with the viral load of the index case: from 12% where the index case had a viral load of <10^6^ copies/mL to 24% where the index case had a viral load >10^9^ copies/mL (Figure 1B). According to the multivariate analysis, the viral load of the index case was strongly associated with the risk of onward transmission (OR per log_10_ increase in VL 1.3; 95% CI 1.1-1.6) (Table 3). Ninety percent (114/125) of transmission events had an index case viral load of 5.1 log_10_ copies/ml or more, and 50% (61/125) had a viral load of 8.8 log_10_ copies/ml or more. Other factors associated with an increased risk of transmission were household contact (OR 2.7, 95% 1.4-5.06) and age of the contact (OR 1.02, 95% 1.01-1.04). There was no association of risk of transmission with reported mask usage by contacts, with the age or gender of the index case nor with the presence of respiratory symptoms in the index case at the initial study visit (Table 3).

**Table 3:**
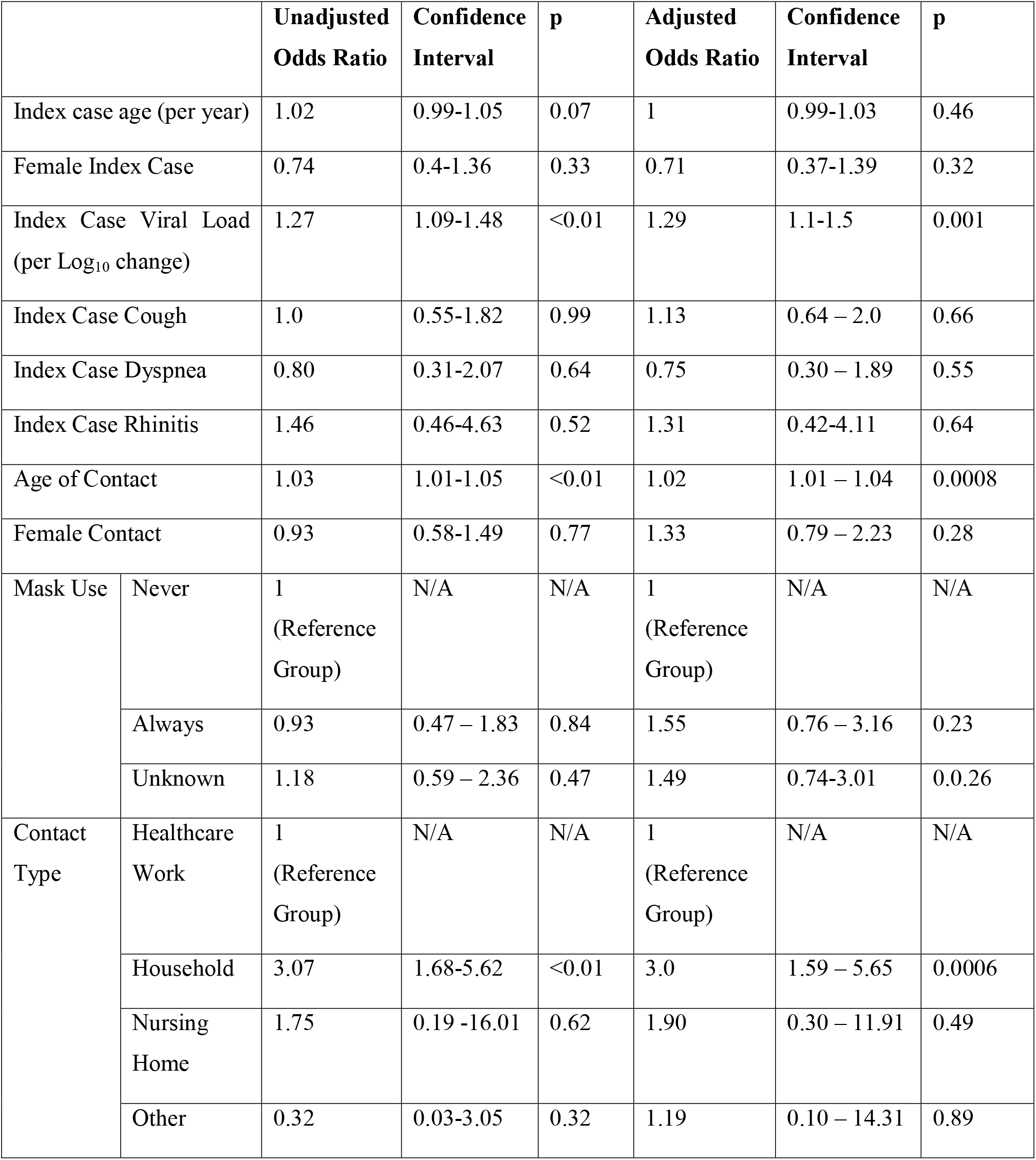
Risk factors for transmission of SARS-CoV-2.

**Figure 1:**
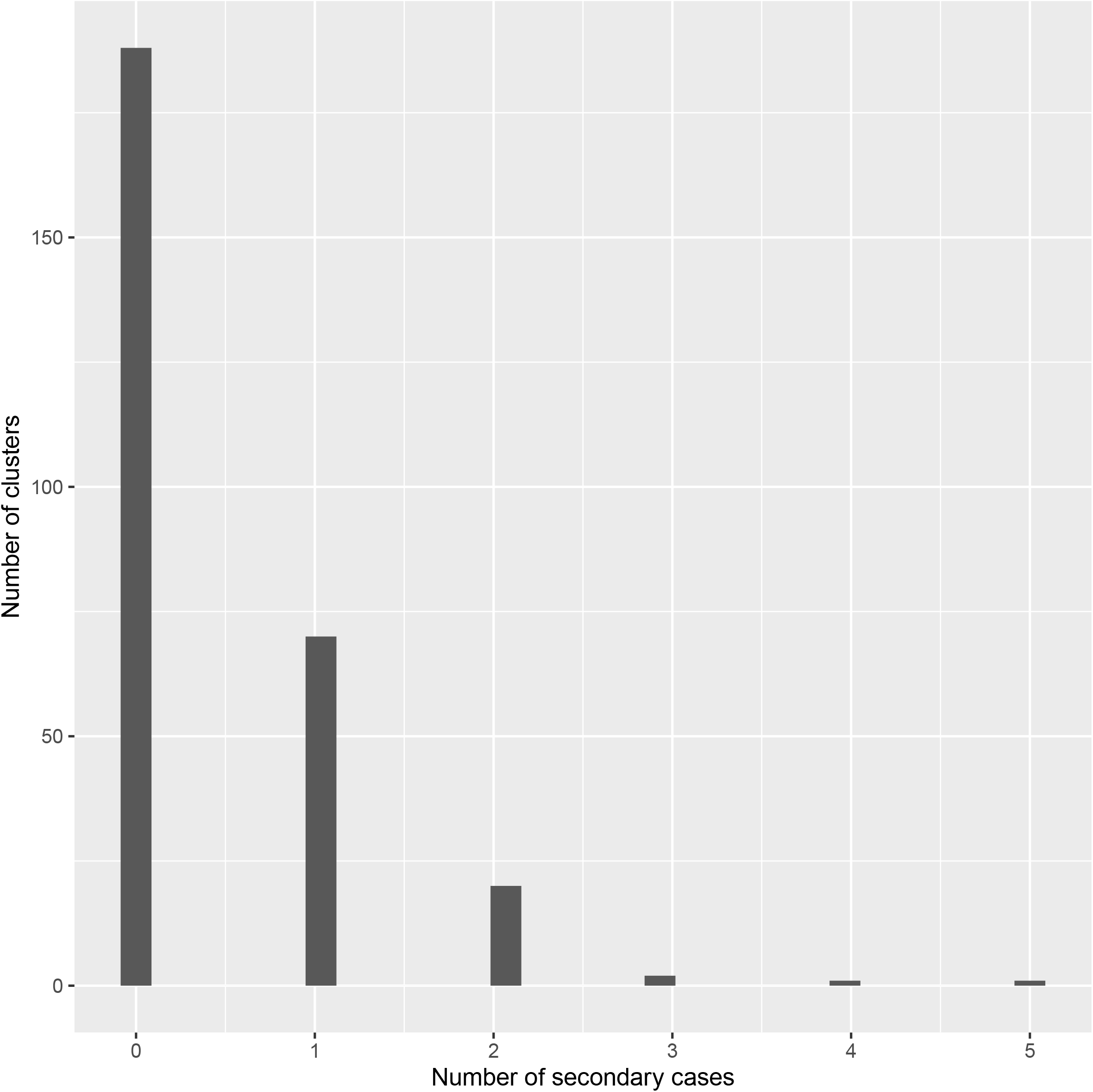
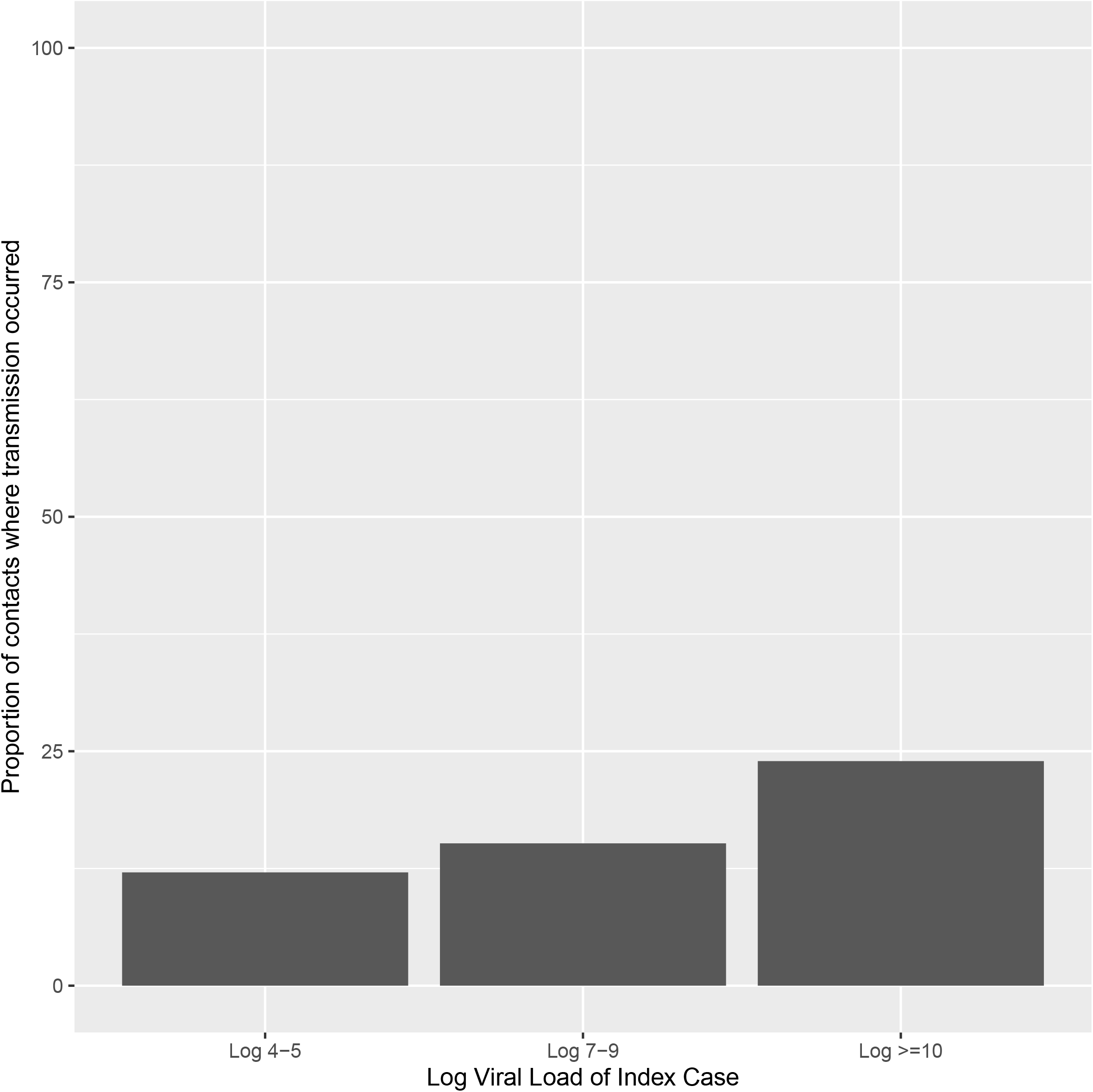
Transmission in a cluster. (A) Number of secondary cases per cluster. (B) Relationship between viral load of the index case and the proportion of contacts developing Covid-19. Numbers 18/149 in group 10^4^-10^5^ RNA copies/ml; 30/2012 in group 10^6^-10^7^; 59/298 in group 10^8^-10^9^; 17/71 in group ≥ 10^10^.

We did not find any evidence of an association between the viral load of the index cases and the first viral load of incident positive results amongst contacts (*p* = 0.1, Supplementary Appendix) and this remained true when adjusting for both the day of illness on which the index cases baseline viral load was measured and the number of days until the contact was enrolled (p = 0.18). Also, after excluding contacts who were PCR positive at the first study visit, we found no association between the viral load of the index case and the time to onset of incident SARS-CoV-2 infection (HR 1.01 95% CI 0.83-1.23).

### Risk factor for Covid-19 disease among PCR+ contacts

Overall, 449 contacts had a positive PCR result at first visit regardless of availability on viral load data of their index case (n=125) or not (n=324). Twenty-eight (6.3%) of 449 contacts had symptoms at the first visit and 181 (40.3%) developed symptomatic Covid-19 within the follow-up period. The multivariable cox-regression analysis, after adjusting for age and sex, revealed that increasing viral load levels of the contact at day 1 were associated with an increased risk of developing symptomatic disease. The risk of symptomatic disease was approximately 25% amongst individuals with an initial viral load of <10^7^ copies/mL compared to a more than 60% amongst those with an initial viral load of >10^9^ (HR per log_10_ increase in VL 1.12; 95% CI 1.05 – 1.2; *p* = 0.0006) (Figure 2A). In the multivariable analysis there was no association between sex or age of individuals nor the presence of diabetes, cardiovascular or respiratory disease and the risk or time to developing symptomatic Covid-19.

**Figure 2.**
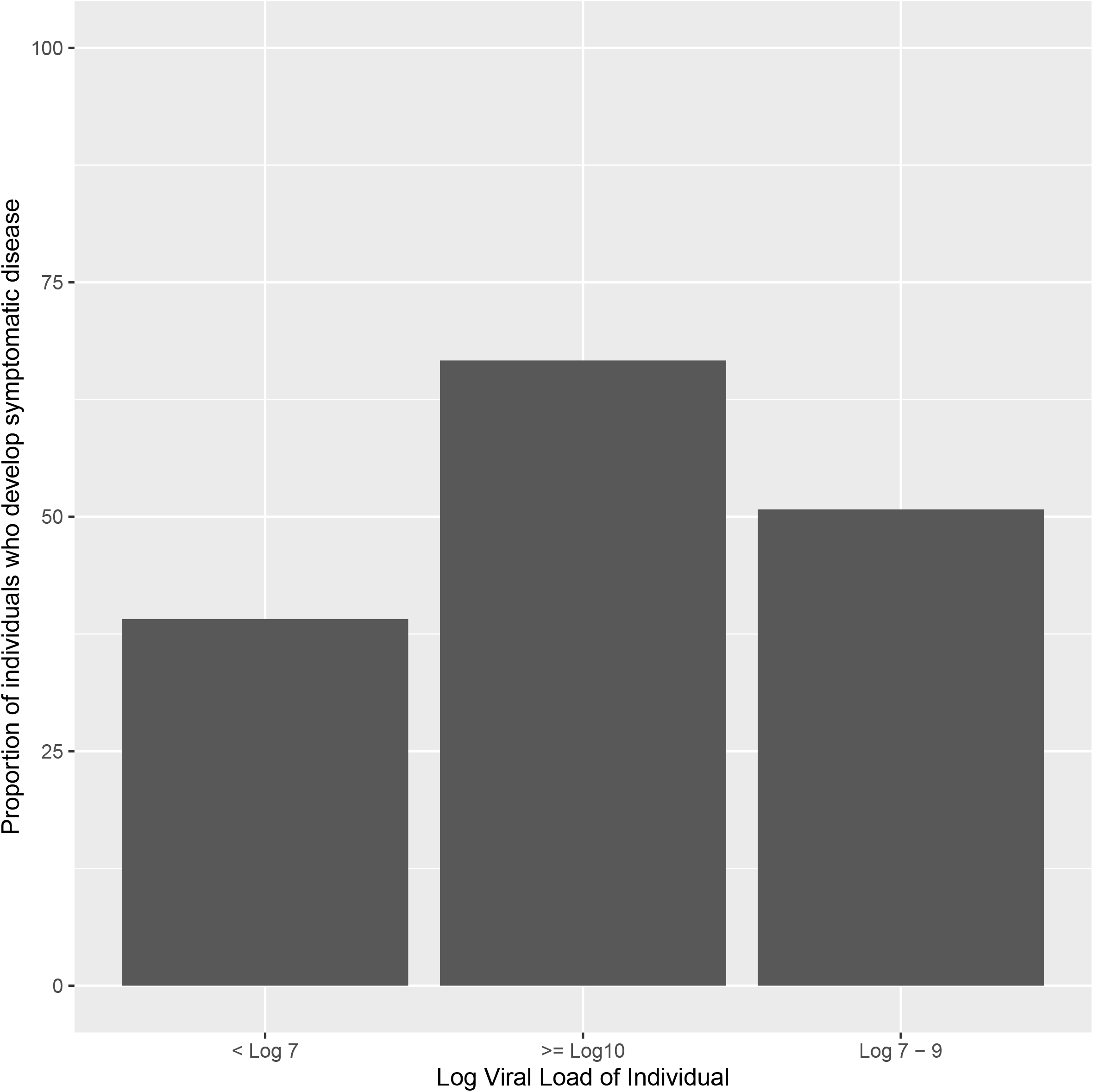
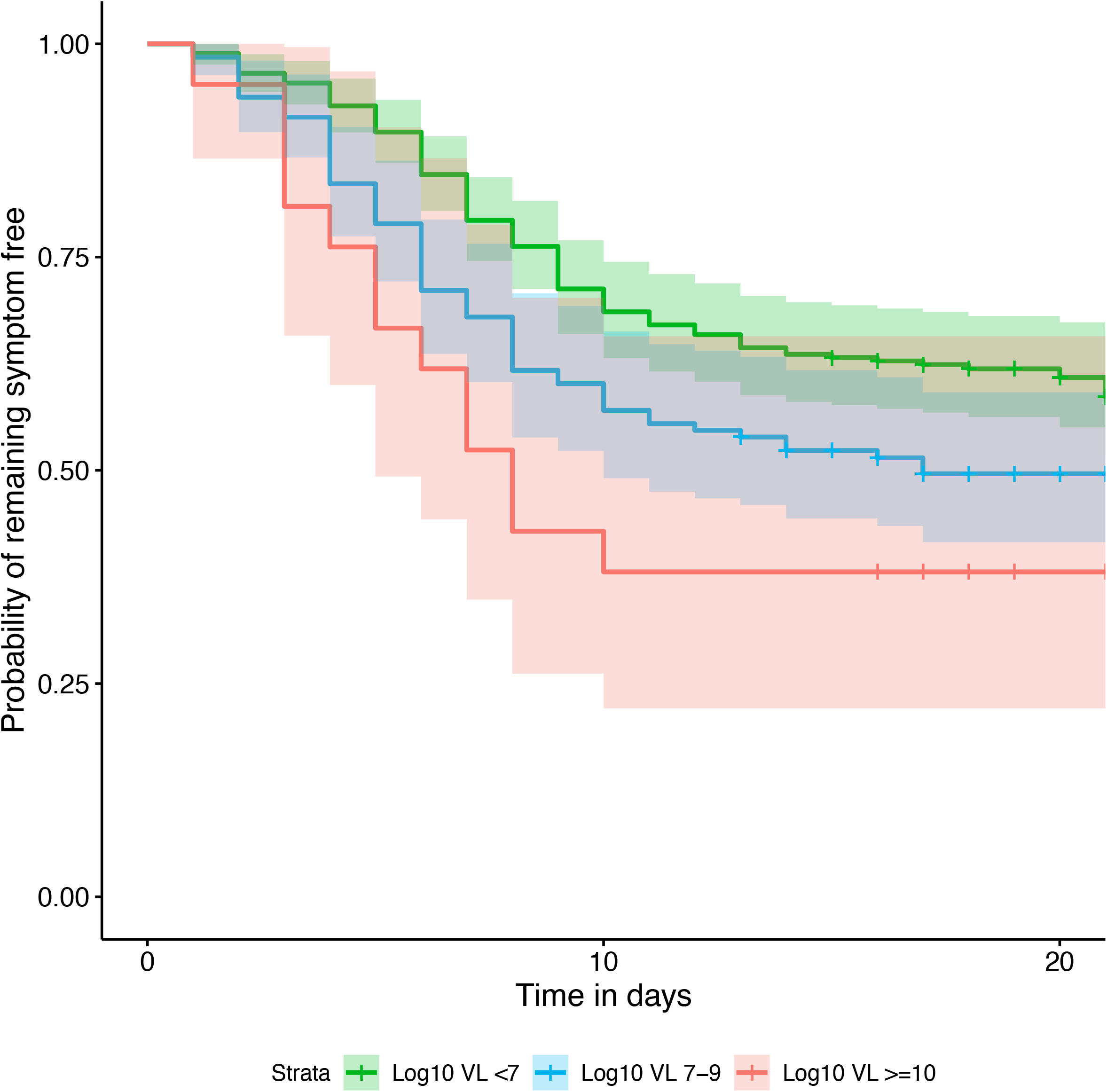
Risk of developing symptomatic Covid-19 according to characteristics of the contact at enrolment. (A) probability of symptomatic disease by viral load. (B) time to symptomatic disease by viral load.

The median time from exposure to symptom onset was 7 days (IQR 5 – 9). The time to onset of symptomatic disease decreased from a median of 7 days (IQR 5 – 10) for individuals with an initial viral load <10^7^ copies/mL to 6 days (IQR 4 – 8) and 5 days (IQR 3 – 8) for individuals with an initial viral load of 10^7^-10^9^ and >10^9^ copies/mL, respectively (Figure 2B). Overall, 110/181 (60.8%) of participants who developed symptoms did so before day 8, 45/181 (24.9%) between days 8-10, and 22/181 (12.2%) between days 11-14.

## DISCUSSION

In our study, we found that increasing viral load values in nasopharyngeal swabs of Covid-19 cases were associated with the greater risk of transmission measured by SARS-CoV-2 PCR positivity among contacts and also a higher risk of transmission in household environment compared to other indoor situations. In addition, we found that higher viral loads in swabs of asymptomatic contacts were associated with higher risk of developing symptomatic Covid-19 and have shorter incubation periods than those with a lower viral load. Relationships between viral load and infectivity have been described for other respiratory viruses and our study confirms the same is true for SARS-CoV-2.

To our knowledge this is the largest study that evaluates the relationship of viral load in Covid-19 cases and risk of transmission. In our cohort, a high proportion (67%) of index cases did not cause secondary infections. However, we identified 90 (33%) clusters with transmission events and the multivariate analysis revealed that clusters centered on index cases with high viral load were significantly more likely to result in transmission. Secondary attack rate was under 12% when the index case viral load was <10^6^ copies/ml compared to more than 20% amongst clusters with the highest viral loads. In line with previous analyses of case-contact clusters,^9,12,14^ we also found that household exposure to an index case was associated with a higher risk of transmission that other types of contact, presumably reflecting duration and proximity of exposure. Age of the contact was also identified in our multivariate analysis as a significant-albeit modest-determinant of transmission. This factor has shown uneven influence across results reported elsewhere, but seems to play a secondary role among adults.^13,14^ Finally, unlike previous analyses that reported a relationship between coughing and transmission,^13^ we did not find any association. This finding suggests that the absence of cough does not preclude significant onward transmission, particularly if the viral load is high. Taken together, our results indicate that the viral load, rather than symptoms, may be the predominant driver of transmission.

Importantly, we report that high viral short after exposure in asymptomatic contacts was strongly associated with the risk of developing symptomatic Covid-19 disease. We found an approximately 25% chance of developing symptomatic disease amongst individuals with an initial viral load <10^7^ copies/mL compared to a more than 60% chance amongst individuals with a viral load >10^9^. These data may provide rationale for risk stratification for developing illness. Moreover, the initial viral load significantly shifted the incubation time, which ranged from 5 days in participants with a high viral load to 7 days in participants with a low viral load. Our study is the first analysis of prospective data that investigates the association between initial viral load and the incubation time.

The study has several limitations. First, asymptomatic people were not enrolled as index cases, affecting our ability to fully characterize all types of transmission chain. Second, we did not find any evidence of decreased risk of transmission in individuals who reported mask use. While this finding collides with the evidence reported elsewhere,^8^ we did not have fine-grained data on type of mask (surgical vs FFP2), use of other measures of PPE or other infection control practices, thus limiting our ability to make clear inferences about the impact of PPE on transmission risk. Mask usage is likely correlated with type of exposure which might further confound associations but we did not note any association between mask use and risk either in our unadjusted analysis (Table 3) or in a multivariable model excluding type of exposure (data not shown). Third, we used time to symptom onset (with later confirmation of infection) rather than time to positive PCR test based on serial testing. Nonetheless, accurate calculation of the incubation period was feasible because of the prospective nature of the study, accurate identification of exposure by face-to-face interview, and intensive active and passive monitoring of exposed contacts. We followed participants over 14-day periods, thus incubation periods beyond 14 days may not have been detected. Within each cluster we cannot be completely certain about the directionality of transmission, but our inclusion criteria including the absence of Covid-19 like symptoms in the 2 weeks proceeding enrolment is consistent with transmission from a case to a contact. We also cannot exclude that some individuals may have been infected by individuals outside of study clusters, but as per national guidelines all contacts were quarantined after exposure to index cases reducing the chance of transmission from elsewhere. Samples were available from index cases a median of four days after symptom onset and the initial sample in contacts was taken on average 5 days after exposure which may limit our ability to detect associations with peak viral load. Despite this we still demonstrate clear dose effects in relation to both risk of transmission and time to symptom onset. Finally, our study population is reflective of the trial from which the study sample is drawn and is therefore biased towards female participants, few comorbidities and predominantly mild-moderate infection and further data is needed on the risk of transmission in other populations.

In summary, our results provide evidence regarding the determinants of SARS-CoV-2 transmission, particularly on the role of the viral load. The higher risk of transmission among individuals with higher viral loads adds to current evidence and encourages assessing viral load in cases with a larger number of close contacts. When a case with high viral load is identified, implementation of reinforced contact tracing measures and quarantines, may be critical to reduce onward transmission. Similarly, our results regarding the risk and expected time to developing symptomatic Covid-19 encourage risk stratification of newly diagnosed SARS-CoV-2 infections based on the initial viral load.

## CONTRIBUTORS

MM, DO, OM accessed and verified the data. MM, DO, ChR, OM conceived of the study. MM performed the analysis. PM, AA, MCM, MU, MVM, CGB, NP, JA, BC, OM led the RCT from which study data is derived. MM and OM wrote the first draft of the manuscript. All authors gave critical input into interpretation and revised the manuscript.

## Data Availability

Data referred to in the manuscript are available upon request to the corresponding author

## ACKNOWLEDGMENTS

The authors would like to thank Gerard Carot-Sans for providing medical writing support during the preparation of the manuscript.

## Data Sharing Statement

Marks M, Millat-Martinez P, Ouchi d et al. Transmission of Covid-19 in 282 clusters in Catalonia, Spain: a cohort study

| Question | Authors' Response |
| --- | --- |
| Will the data collected for your study be made available to others? | Yes |
| Which data? | Complete de-identified patient data set |
| Will any supporting documents be available? | Study protocol |
| How or where can the data be obtained? |  |
| When will data availability begin? | 15/12/2020 |
| When will data availability end? | 15/12/2021 |
| To whom will data be available? | Researchers whose proposed use of the data has been approved |
| For what type of analysis or purpose? | For any purpose |
| By what mechanism? | By email |
| Any other restrictions? | — |
| Additional information | — |

## REFERENCES

1 La Rosa G, Bonadonna L, Lucentini L, Kenmoe S, Suffredini E. Coronavirus in water environments: Occurrence, persistence and concentration methods - A scoping review. Water Res 2020; 179: 115899.

2 Umakanthan S, Sahu P, Ranade A V., et al. Origin, transmission, diagnosis and management of coronavirus disease 2019 (COVID-19). Postgrad Med J 2020; 0: 1–6.

3 Leclerc QJ, Fuller NM, Knight LE, Funk S, Knight GM. What settings have been linked to SARS-CoV-2 transmission clusters? Wellcome Open Res 2020; 5: 83.

4 Qian H, Miao T, Liu L, Zheng X, Luo D, Li Y. Indoor transmission of SARS-CoV-2. medRxiv 2020;: 2020.04.04.20053058.

5 Hamner L, Dubbel P, Capron I, et al. High SARS-CoV-2 Attack Rate Following Exposure at a Choir Practice — Skagit County, Washington, March 2020. MMWR Morb Mortal Wkly Rep 2020; 69: 606–10.

6 Park SY, Kim YM, Yi S, et al. Coronavirus disease outbreak in call center, South Korea. Emerg Infect Dis 2020; 26: 1666–70.

7 Muñoz MA, López-Grau M. Lessons learned from the approach to the COVID-19 pandemic in urban primary health care centres in Barcelona, Spain. Eur J Gen Pract 2020; 26: 106–7.

8 Chu DK, Akl EA, Duda S, et al. Physical distancing, face masks, and eye protection to prevent person-to-person transmission of SARS-CoV-2 and COVID-19: a systematic review and meta-analysis. The Lancet 2020; 395: 1973–87.

9 Bi Q, Wu Y, Mei S, et al. Epidemiology and transmission of COVID-19 in 391 cases and 1286 of their close contacts in Shenzhen, China: a retrospective cohort study. Lancet Infect Dis 2020; 20: 911–9.

10 Wölfel R, Corman VM, Guggemos W, et al. Virological assessment of hospitalized patients with COVID-2019. Nature 2020; 581: 465–9.

11 La Scola B, Le Bideau M, Andreani J, et al. Viral RNA load as determined by cell culture as a management tool for discharge of SARS-CoV-2 patients from infectious disease wards. Eur J Clin Microbiol Infect Dis 2020; 39: 1059–61.

12 Böhmer MM, Buchholz U, Corman VM, et al. Investigation of a COVID-19 outbreak in Germany resulting from a single travel-associated primary case: a case series. Lancet Infect Dis 2020; 20: 920–8.

13 Wu J, Huang Y, Tu C, et al. Household Transmission of SARS-CoV-2, Zhuhai, China, 2020. Clin Infect Dis 2020; published online May 11. DOI:10.1093/cid/ciaa557.

14 Cheng HY, Jian SW, Liu DP, Ng TC, Huang WT, Lin HH. Contact Tracing Assessment of COVID-19 Transmission Dynamics in Taiwan and Risk at Different Exposure Periods before and after Symptom Onset. JAMA Intern Med 2020; 180: 1156–63.

15 Huang L, Zhang X, Zhang X, et al. Rapid asymptomatic transmission of COVID-19 during the incubation period demonstrating strong infectivity in a cluster of youngsters aged 16-23 years outside Wuhan and characteristics of young patients with COVID-19: A prospective contact-tracing study. J Infect 2020; 80: e1–13.

16 Liu T, Gong D, Xiao J, et al. Cluster infections play important roles in the rapid evolution of COVID-19 transmission: a systematic review. Int J Infect Dis 2020; 99: 374.

17 Buitrago-Garcia D, Egli-Gany D, Counotte MJ, et al. Occurrence and transmission potential of asymptomatic and presymptomatic SARS-CoV-2 infections: A living systematic review and meta-analysis. PLOS Med 2020; 17: e1003346.

18 Backer JA, Klinkenberg D, Wallinga J. Incubation period of 2019 novel coronavirus (2019-nCoV) infections among travellers from Wuhan, China, 20 28 January 2020. Eurosurveillance 2020; 25: 20–8.

19 Li Q, Guan X, Wu P, et al. Early transmission dynamics in Wuhan, China, of novel coronavirus-infected pneumonia. N Engl J Med 2020; 382: 1199–207.

20 Leung C. The difference in the incubation period of 2019 novel coronavirus (SARS-CoV-2) infection between travelers to Hubei and nontravelers: The need for a longer quarantine period. Infect Control Hosp Epidemiol 2020; 41: 594–6.

21 Mitja O, Ubals M, Corbacho M, et al. A Cluster-Randomized Trial of Hydroxychloroquine as Prevention of Covid-19 Transmission and Disease. medRxiv 2020;: 2020.07.20.20157651.

22 Catalan Ministry of Health. Catalan epidemiological surveillance system. http://salutpublica.gencat.cat/ca/ambits/vigilancia_salut_publica/ (accessed March 28, 2020).

23 Centers for Disease Control and Prevention (CDC). CDC 2019-Novel Coronavirus (2019-nCoV) Real-Time RT-PCR Diagnostic Panel. Cat. 2019-NCoVEUA-01. 2020. https://www.fda.gov/media/134922/download (accessed May 21, 2020).

24 World Health Organization. The First Few X (FFX) Cases and contact investigation protocol for 2019-novel coronavirus (2019-nCoV) infection, version 2. https://www.who.int/publications/i/item/the-first-few-x-(ffx)-cases-and-contact-investigation-protocol-for-2019-novel-coronavirus-(2019-ncov)-infection (accessed Sept 21, 2020).

