## Supplementary figures and images for "Transmission of COVID-19 in 282 clusters in Catalonia, Spain: a cohort study"

### Supplementary Figure 1

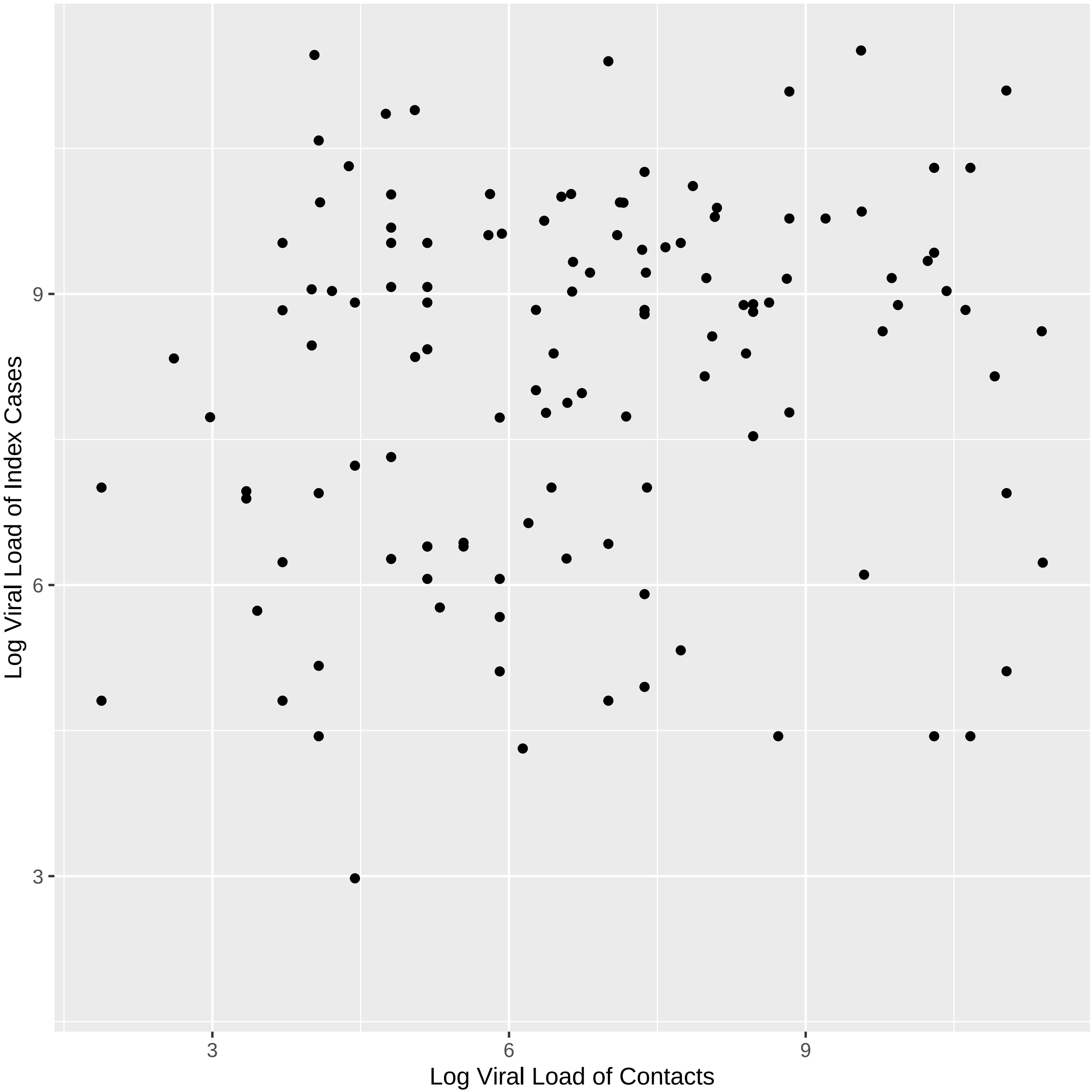
