## Supplementary Material for "Transmission of COVID-19 in 282 clusters in Catalonia, Spain: a cohort study"

**PCR:**
Serial SARS-CoV-2 PCR test and viral load titration on nasopharyngeal swab were conducted on day 1 and day 14 to all participants, and on any unscheduled visit when the participant notified the onset of Covid-19 symptoms. The detection of the SARS-CoV-2 virus was performed from nasopharyngeal swabs at SYNLAB Diagnostics (Barcelona, Spain) by PCR using TaqMan™ 2019-nCoV Assay Kit according to the manufacturer's protocol (Catalog number: A47532, Thermo Fischer Scientific Inc.). Viral load was quantified from nasopharyngeal swabs at IrsiCaixa laboratory (Badalona, Spain) by PCR amplification, based on the 2019-Novel Coronavirus Real-Time RT-PCR Diagnostic Panel guidelines and protocol developed by the American Center for Disease Control and Prevention (CDC).[22] For absolute quantification, a standard curve was built using 1/5 serial dilutions of a SARS-CoV2 plasmid (2019-nCoV_N_Positive Control, catalog no. 10006625, $2x10^5$ copies/µL, Integrated DNA Technologies) and run in parallel to all PCR determinations.

Approximate relationships between CT values and quantitative viral load are shown below:

| Log Viral Load | Cycle Threshold |
|---|---|
| $10^4$ | 35-37 |
| $10^6$ | 29-31 |
| $10^8$ | 23-25 |
| $10^{10}$ | 17-19 |

Proportional Hazards Assumption for Cox Regression

We assessed the proportional hazards assumption by assessing scaled Schoenfeld residuals. There was no association between residuals and time for any of the covariates nor globally confirming that the proportional hazards assumption was met.

| Variable | P |
|---|---|
| Viral Load | 0.21 |
| Age | 0.75 |
| Sex | 0.17 |
| Global | 0.33 |

Graphical demonstrations are given below confirming the proportional hazards assumptions holds in Figure 1.

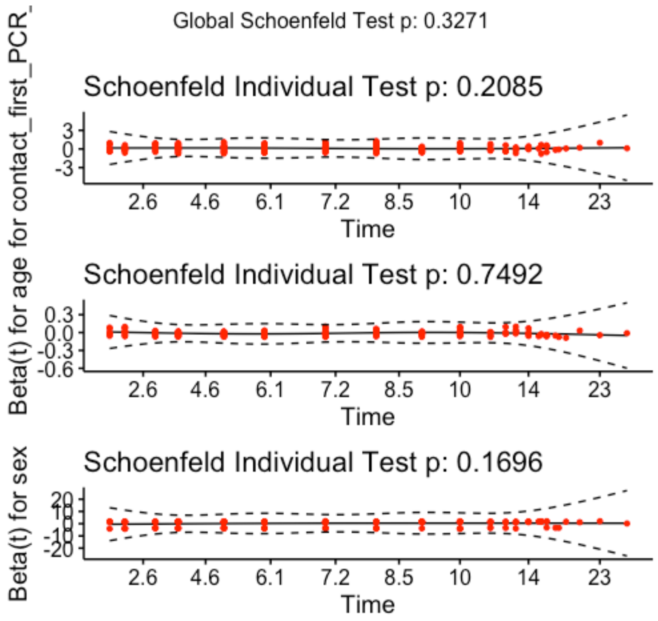


**Supplementary Figure 2: Relationship between Index Case and Contacts Viral Load**

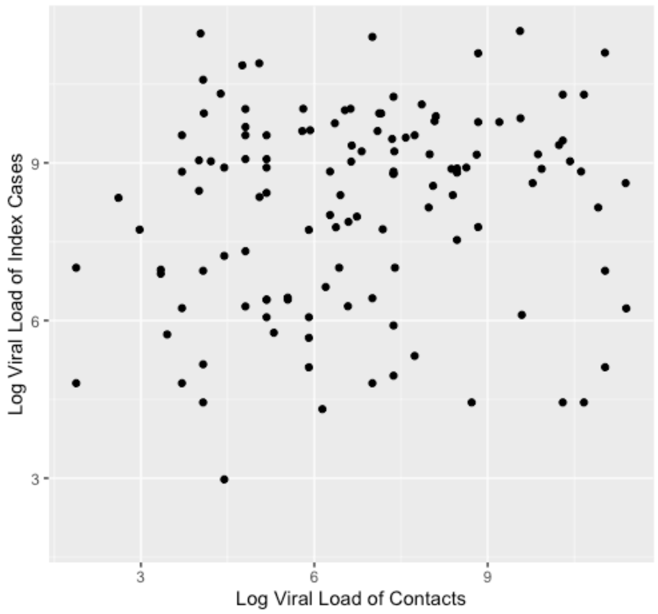
